# Early assessment of potential airline-mediated importation risk during the 2026 DRC-Uganda Bundibugyo virus disease outbreak

**DOI:** 10.64898/2026.06.01.26354569

**Authors:** Ryo Kinoshita, Motoi Suzuki, Daisuke Yoneoka

## Abstract

During the 2026 Bundibugyo virus disease outbreak in the Democratic Republic of the Congo and Uganda, we projected potential airline-mediated importation risk using contemporary airline network and an externally calibrated Ebola importation hazard. Effective-distance analyses identified major international hub countries, including Belgium, France, South Africa, Kenya, and the United Arab Emirates, as higher-probability gateways until the end of June, 2026. These early projections provide a reproducible framework for real-time international situational awareness, while emphasizing that importation risk does not imply local transmission risk.

---

On 15 May 2026, an outbreak of Bundibugyo virus disease (BVD) was reported in the Democratic Republic of the Congo (DRC), with cross-border importations subsequently detected in Uganda. On 17 May 2026, the World Health Organization (WHO) declared the event a Public Health Emergency of International Concern (PHEIC) [1]. BVD is caused by *Bundibugyo virus*, an orthoebolaviruses that cause Ebola disease (EBOD). As of 4 June 2026, the DRC had reported 363 confirmed cases, including 62 deaths [1–3]. Uganda had reported 15 confirmed cases, including one death. Although no airline-mediated importation had been documented for the current outbreak, international air travel remains a plausible route for long-distance dissemination if exposed individuals travel during the incubation period or early phase of illness. We projected the potential airline-mediated importation risk during the 2026 DRC-Uganda outbreak using a hazard-based model informed by contemporary airline transportation networks and a hazard parameter calibrated from the 2013–2016 West African Ebola virus disease (EVD) epidemic, one of the few EBOD outbreaks with documented long-distance airline-mediated international exportations.

## Airline-flow-based network and effective distance for hazard estimation

To model the global spread on airline network, we first constructed a directed, weighted air-traffic network using airport-level origin–destination passenger-flow data from OAG for May 2024. OAG is a global travel data provider listed in the IATA Strategic Partners Directory and provides aviation schedules, status data, and analytics products for the air-travel ecosystem. The network includes 11,264 airports (nodes), 569,741 routes (edges) and 329,326,078 passengers. Given a single airport-level adjacency matrix *A*, where *A*_*ij*_ denotes the number of passenger volume travelling from airport *i* to *j* during May 2024, the shortest-path effective distance from Kinshasa/N’djili International Airport (IATA code: FIH) to airport *j* was defined by 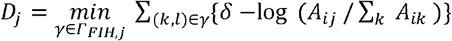, where *Γ*_*ij*_ indicates the set of all directed paths from airport *i* to *j* and *γ* is one path. Shortest paths were calculated on the directed network using Dijkstra’s algorithm. In the main analyses, disease-specific values of *δ* was set to 1, which assumes that the each additional travel step contributes one unit of distance, and thus corresponds to the Brockmann–Helbing scale [4]. This scale has been empirically shown to effectively predict the global spread patterns of infectious diseases [4–7]. In the sensitivity analysis, *δ* can be also estimated from 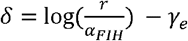 [8], where *r* is the epidemic growth rate (*r* = {0.034, 0.05, 0.1} estimated as of May 17, 2026 [9], *γ_e_* is the Euler–Mascheroni constant (*γ*_e_ ≈ 0.58), and *α*_FIH_ is the outbound passenger flow from FIH divided by the assumed FIH catchment population *α*_FIH_ ≈ 0.0001. While this *δ* formulation allows us to incorporate information about the current epidemic situation by adding more parameters, it also increases uncertainty.

Then, to estimate the arrival probability using the effective distance, we modeled the arrival hazard at airport *j* as *λ*_*j*_ = θ/*D_j_*, where *θ* = 0.19 is used as the primary value based on a previous EBOD importation hazard model [7]. The arrival probability that infection reaches airport *j* within *t* days is calculated by *p_j_*(*t*) = 1 - exp(-*tλ_j_*). As the sensitivity analysis, we checked *θ* = {0.06, 0.32} based on the 95% confidence interval of [7]. Further, the country-level arrival risk was summarized as the maximum airport-level probability within each country: 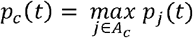, where *A_c_* denotes the set of airports located in country *c*. Lastly, we performed the same analysis using Entebbe International Airport (IATA code: EBB) as the starting point instead of FIH.

### Estimated importation risk

Figure 1 shows the airline passenger-flow network flying from FIH in DRC and their estimated effective distance. Effective-distance pathways were shaped by airline connectivity rather than geographic distance alone, with highly connected transit airports serving as major intermediate nodes. The estimated effective distance ranged from 3.02 to 29.5 (mean of 15.41).

**Figure 1.**
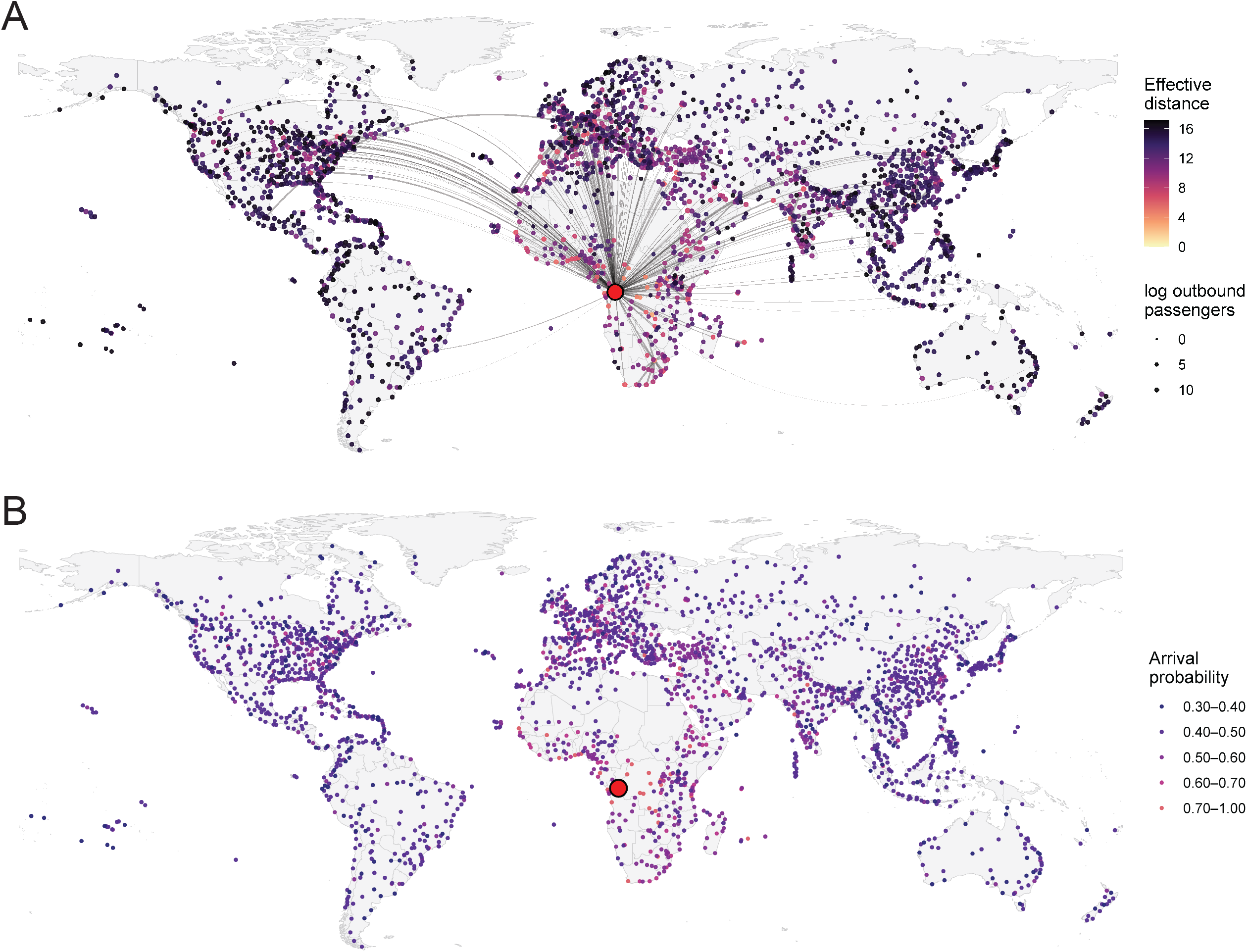
(A) Shortest-path effective distance from Kinshasa/N’djili International Airport (IATA code: FIH) on the airline-flow network. (B) Estimated arrival probability from Kinshasa/N’djili International Airport (IATA code: FIH) by the end of June with θ=0.19 and δ=1.

Airport-level arrival probabilities, interpreted as the probability of at least one airline-mediated importation until the end of June, 2026 (*t* = 44), varied substantially across the global airline network (Figure 2). The probability ranged from 0.247 to 0.937 (mean of 0.431). Higher arrival probabilities were concentrated among airports with shorter effective distances from FIH and stronger connectivity through major transit hubs: for example, 0.889 in Brussels Airport, 0.888 in Paris Charles de Gaulle Airport, and 0.871 in Johannesburg O.R. Tambo International Airport (Supplementary Table 1). The sensitivity analysis shows that the probability had a similar geographical tendency, even if the starting airport is assumed to be EBB: the mean (standard deviation) ranged from 0.170 (0.049) to 0.616 (0.099) (Supplementary Figures 1-5 and Supplementary Table 2).

Table 1 shows the top-10 country-level arrival probability ranked by *p_c_*(*t*). The highest 30-day country-level arrival probabilities were observed in major international hub countries including Belgium, France, South Africa, Kenya, and the United Arab Emirates, with the top-ranked countries all exceeding 0.77, whose probaility ranged from 0.38 to 0.98 in the sensitivity analysis.

**Table 1.** Top-10 country-level arrival probability ranked by *p_c_*(*t*).

| Country | Number of Airports | Effective distance (Mean) | Maximum airport arrival probability, $p_c(t)$ | | |
| --- | --- | --- | --- | --- | --- |
| | | | $\theta = 0.19$ | $\theta = 0.06$ | $\theta = 0.32$ |
| Belgium | 5 | 12.373 | 0.889 | 0.501 | 0.975 |
| France | 45 | 12.552 | 0.889 | 0.499 | 0.975 |
| South Africa | 21 | 9.543 | 0.871 | 0.477 | 0.968 |
| Kenya | 23 | 12.352 | 0.862 | 0.465 | 0.964 |
| United Arab Emirates | 7 | 10.929 | 0.846 | 0.446 | 0.957 |
| Türkiye | 51 | 11.071 | 0.827 | 0.426 | 0.948 |
| Uganda | 6 | 10.094 | 0.822 | 0.420 | 0.945 |
| India | 110 | 13.117 | 0.814 | 0.412 | 0.941 |
| Cote d'Ivoire | 6 | 10.467 | 0.784 | 0.384 | 0.925 |
| Canada | 211 | 16.674 | 0.778 | 0.379 | 0.921 |

### Interpretation and limitations

We estimated potential global BVD importation risk during the early phase using a large airline-network dataset comprising over 300 million passenger volumes. Because no airline-mediated importation had been reported so far as of 28 May 2026, we used a hazard parameter estimated using the 2013–2016 West African EVD epidemic [7]. Here, we assume that the relationship between effective distance and EVD importation observed in 2013–2016 was transferable to the current BVD outbreak. The estimated effective distance distribution was very similar to that in the previous study [7], supporting the use of same *θ* value on a comparable hazard scale. Given the limited early outbreak data, this approach can provide a reproducible framework for translating historical importation experience and contemporary airline connectivity into early situational awareness.

Sensitivity analyses using the lower and upper bounds of the historical parameter changed absolute arrival probabilities but broadly preserved the overall pattern of higher-probability airports. This is expected because effective distance determines the spatial pattern, whereas the hazard parameter mainly scales the probabilies. Our estimates also relied on 2024 airline network data and therefore may not capture subsequent changes in routes, passenger volumes, airspace restrictions, or travel advisories, including those affecting routes through Russia and the Middle East. The projections also did not explicitly account for post-PHEIC interventions, enhanced screening or behavioural changes. More importantly, our analysis focused on global air travel originationg from FIH, with EBB considered in sensitivity analysis, and did not explicitly account for land-based spread within the DRC or cross-border movements to neighboring countries.

Estimated arrival probability should be interpreted as importation risk rather than the risk of local transmission after arrival. Importation does not necessarily lead to onward spread because EBOD requires direct contact with infectious body fluids or contaminated materials, and timely detection, isolation, contact tracing and infection prevention and control can reduce transmission [10]. Thus, countries with high-probability airport gateways should be interpreted as priorities for awareness and preparedness, rather than as locations where local outbreaks are expected.

## Conclusion

Our analysis provides an early, and reproducible assessment of potential long-distance dissemination pathways. These projections can support timely international situational awareness and should be updated as additional epidemiological information becomes available, including reports of exported cases.

## Table and figure legends

## Supplementary table and supplementary figure legends

Supplementary table 1. Top-100 airport-level arrival probability from Kinshasa/N’djili International Airport (IATA code: FIH)

Supplementary table 2. Top-100 airport-level arrival probability from Entebbe International Airport (IATA code: EBB)

Supplementary figure 1. Sensitivity analysis of *θ*

Supplementary figure 2. Sensitivity analysis using *r*=0.034

Supplementary figure 3. Sensitivity analysis using *r*=0.05

Supplementary figure 4. Sensitivity analysis using *r*=0.1

Supplementary figure 5. Estimated arrival probability from Entebbe International Airport (IATA code: EBB) by the end of June with δ=1.

## Contributors

RK and DY led the study. All authors took responsibility for the integrity of the data and the accuracy of the data analysis. All the authors made critical revisions to the manuscript for important intellectual content and gave final approval of the manuscript. The opinions, results, and conclusions reported in this paper are those of the authors and are independent from the funding bodies.

## Data availability

The airline passenger-flow data used in this study were obtained from OAG under a commercial licence and cannot be shared publicly by the authors because of third-party data-use restrictions. Researchers interested in accessing these data should contact OAG directly. The epidemiological data used to contextualise the outbreak were obtained from publicly available sources cited in the manuscript. Derived aggregate results supporting the findings of this study are available within the article and its supplementary material.

## Collaborators

N/A

## Ethical statement

This study was based on publicly available aggregate outbreak data and aggregate airline transportation data. It did not involve human participants, individual-level records, or identifiable personal information; therefore, ethical approval and informed consent were not required.

## Use of artificial intelligence tools

During the preparation of this work, the authors used ChatGPT for English language editing and wording refinement. The authors reviewed and edited the output as needed and take full responsibility for the content of the published article.

## Preprint

None at the time of submission.

## Acknowledgements

None

## Other notes/disclaimers

None

