## Supplementary figures and images for "Early assessment of potential airline-mediated importation risk during the 2026 DRC-Uganda Bundibugyo virus disease outbreak"

### Supplementary figure 1

#### Arrival probability from FIH after 44 days

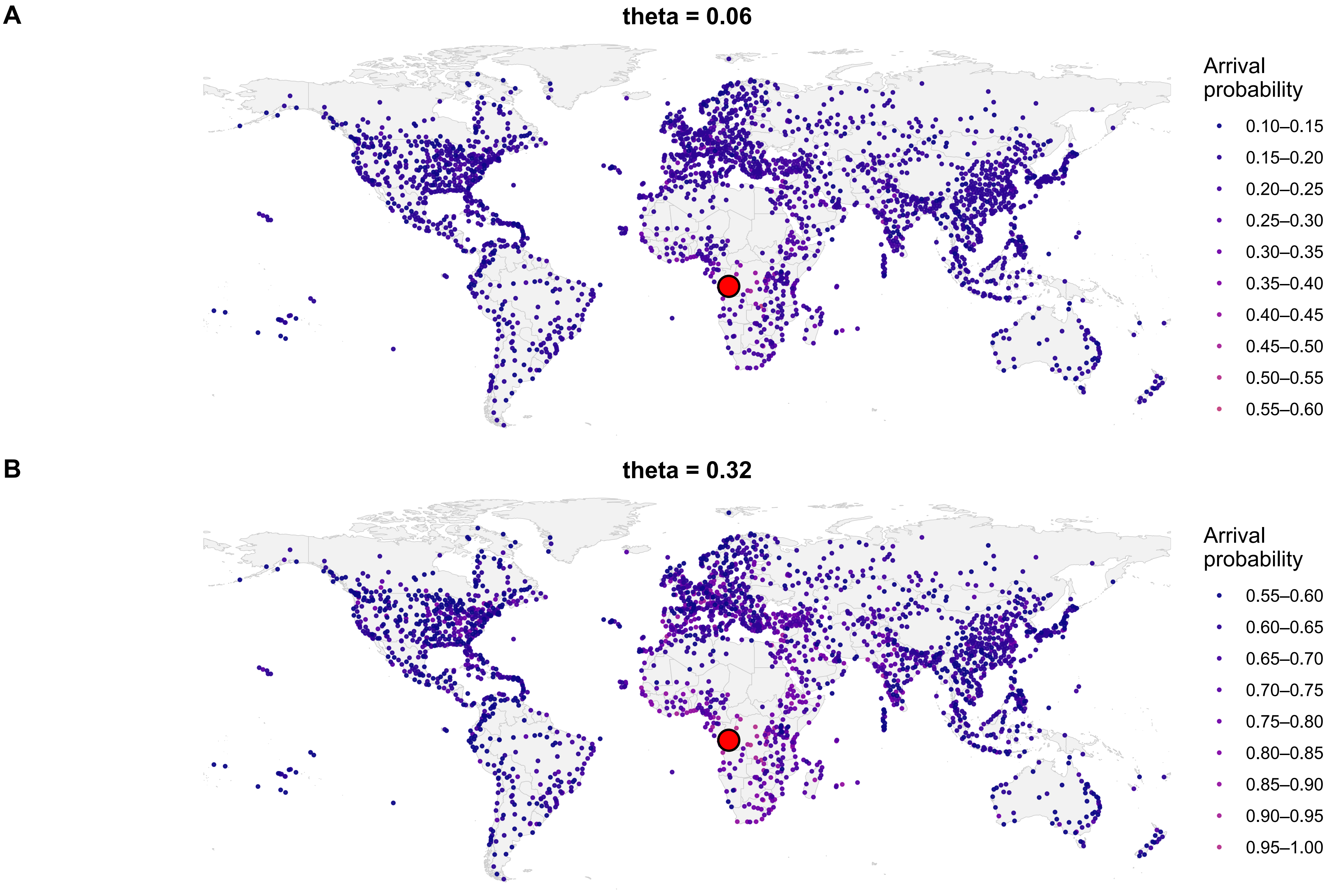

### Supplementary figure 2

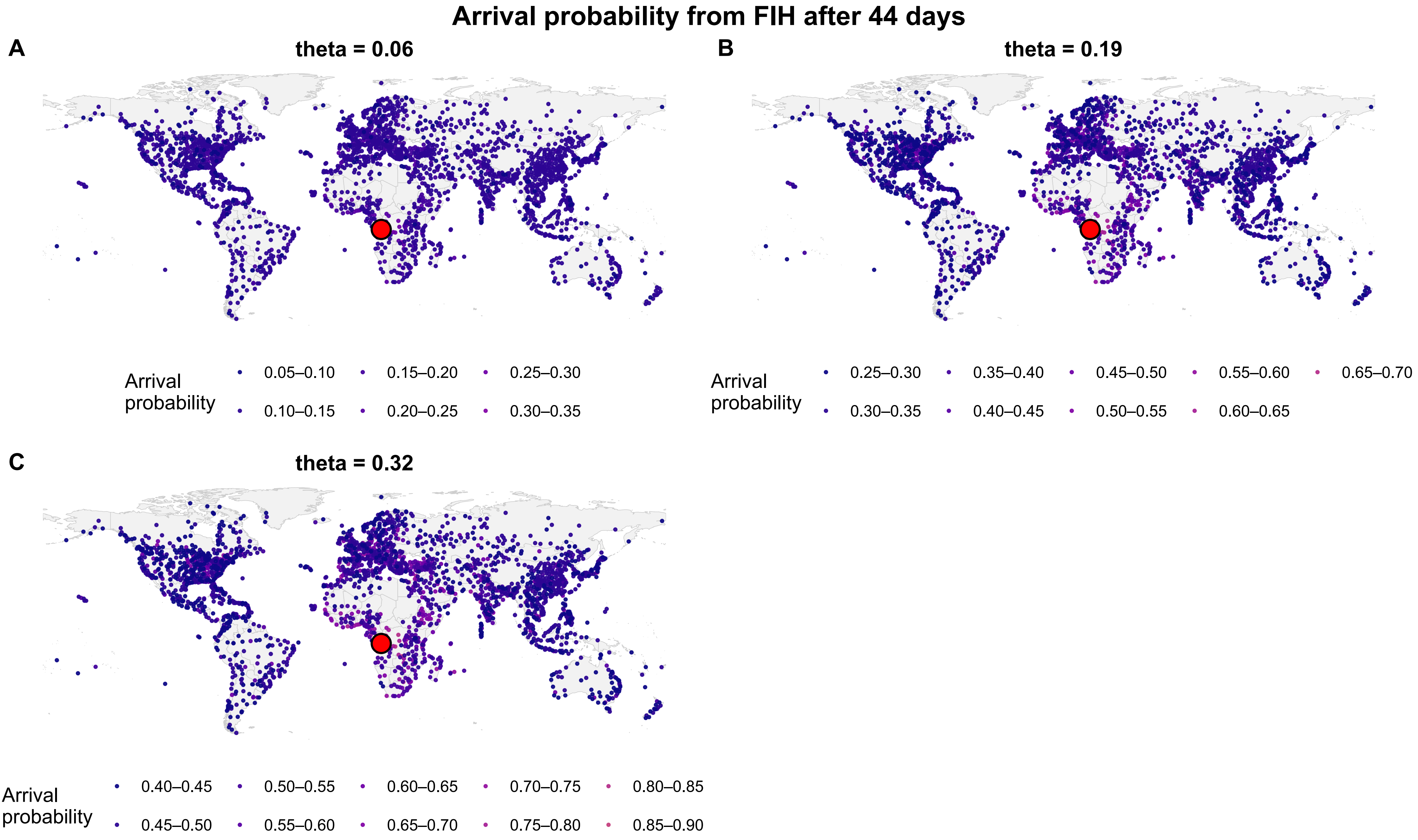

### Supplementary figure 3

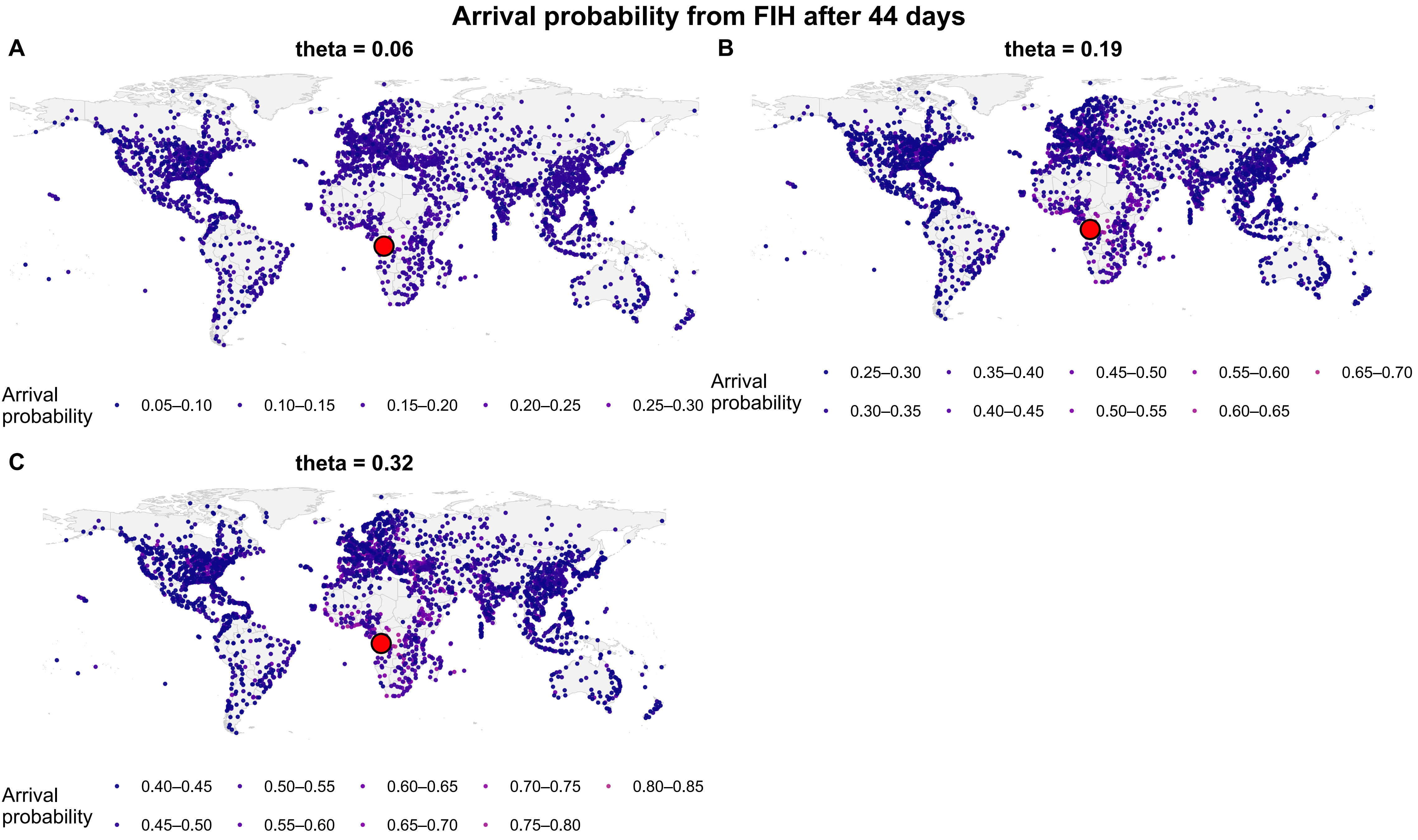

### Supplementary figure 4

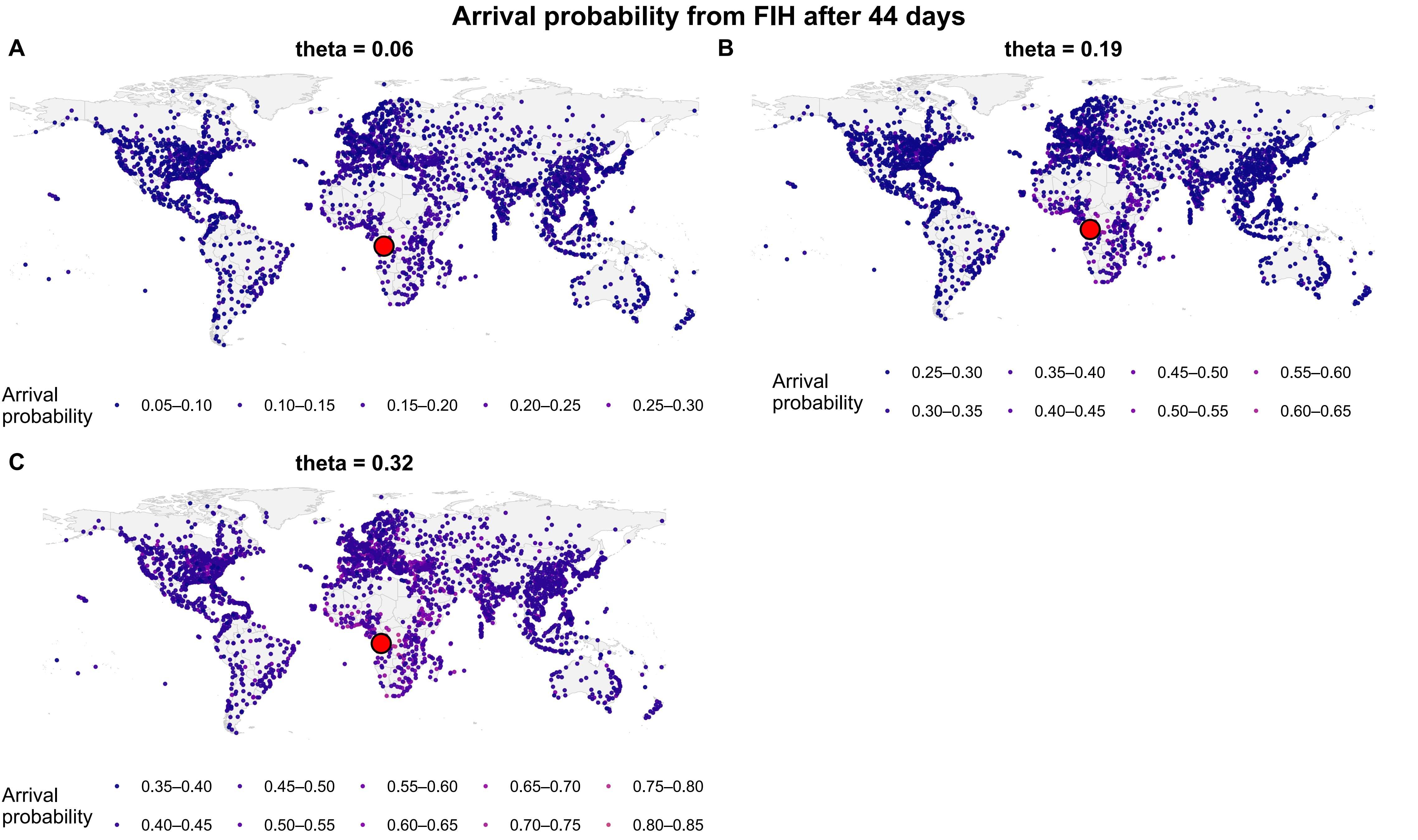

### Supplementary figure 5

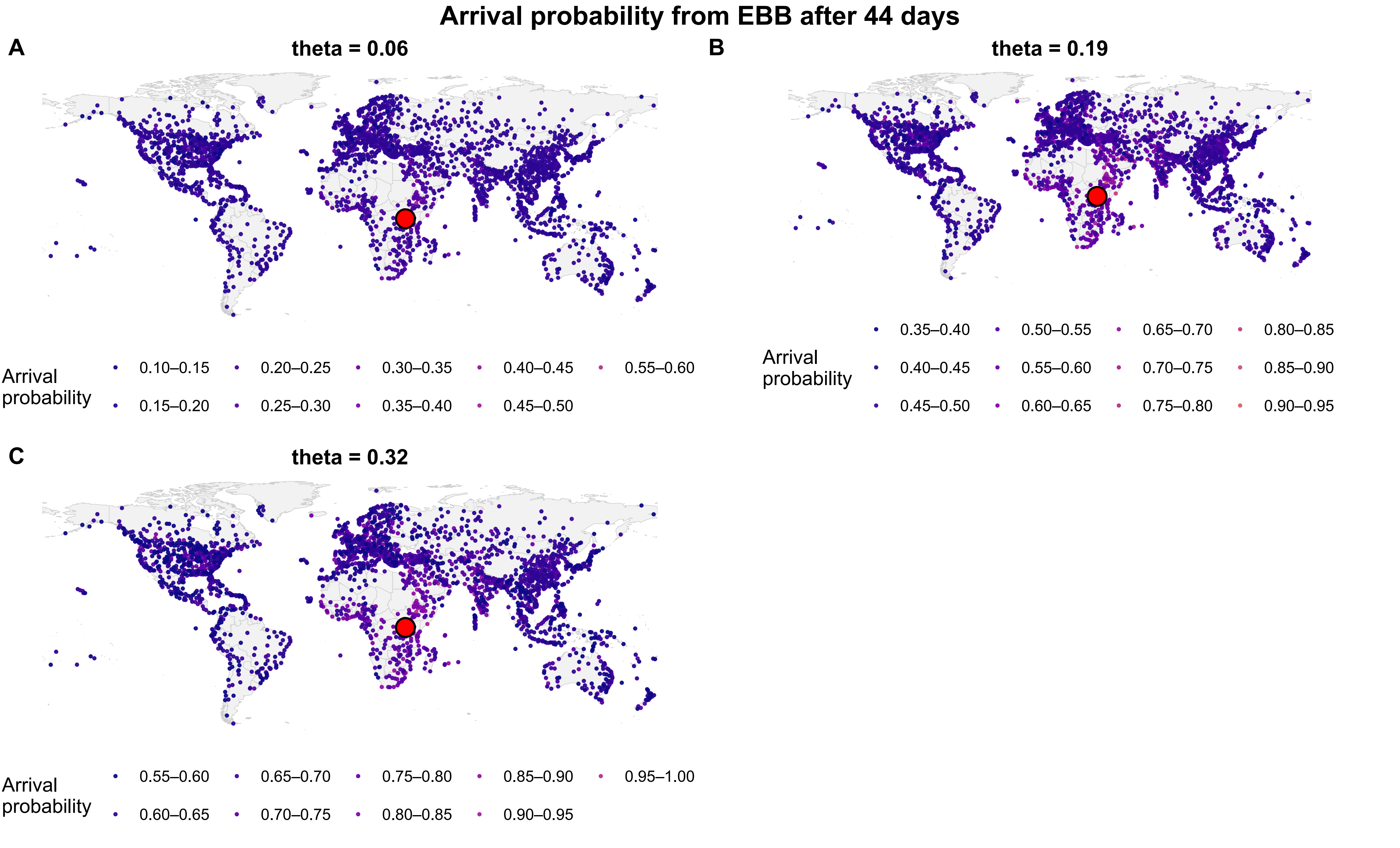
